# Effect of Airborne Particulate Matter on mtDNA Copy Number: A Systematic Review and Meta- Analysis

**DOI:** 10.64898/2026.08.20.26360559

**Authors:** Ashwani Pathak, Akansha Tandekar, Ashutosh Kumar Singh, Vikas Gurjar, Devojit Kumar Sarma, Ram Kumar Nema, Rajnarayan Tiwari, Pradyumna Kumar Mishra

## Abstract

Ambient particulate matter (PM) is a well-established environmental risk factor for non-communicable diseases, yet its influence on mitochondrial function remains poorly defined. Mitochondrial DNA copy number (mtDNA-CN) serves as a biomarker of mitochondrial biogenesis and is a candidate exposure biomarker. We conducted the study in line with PRISMA guidelines (PROSPERO-CRD420261320957) and examined the association between ambient PM exposure and mtDNA-CN. We assessed the risk of bias using Joanna Briggs Institute tools and pooled relative and absolute changes in mtDNA-CN using random-effects models, with subgroup analyses by pollutant type and a descriptive synthesis of mechanistic evidence. Of 1,224 records identified, 24 studies met inclusion criteria for quantitative analysis, with 12 reporting percentage change and 12 reporting absolute values, covering 13,092 participants. PM exposure was significantly associated with decreased percentage mtDNA-CN (ES: -4.90; 95% CI: -7.97 to -1.82; p = 0.002), while absolute mtDNA-CN levels increased significantly (ES = 0.55; 95% CI: 0.05 to 1.04; p = 0.030). Contributing mechanistic pathways included mtDNA hypermethylation, impaired mitochondrial biogenesis, and mitochondrial dynamics. Our findings show that ambient PM exposure alters mtDNA-CN, though the directionality differs across metrics, highlighting the need for larger prospective studies to validate mtDNA-CN as a reliable biomarker of airborne PM and nanoparticulate exposure.

## Introduction

Air pollution is one of the biggest environmental threats to human health worldwide and is linked to roughly 4.2 million premature deaths every year (Cohen et al., 2017; Fuller et al., 2022). Particulate matter (PM) is a big part of that problem, and its physical and chemical makeup varies so much that its harm varies right along with it (Kwon et al., 2020). Studies have linked PM exposure to heart and lung disease, metabolic problems, cognitive decline, age-related illness, and early death (Schraufnagel et al., 2018). PM is grouped into coarse (PM₁₀), fine (PM₂.₅), and ultrafine (PM₀.₁) categories by particle size; its chemical makeup and source also shape its toxicity (Kelly et al., 2015). Components include PAHs, diesel exhaust particles, and black carbon, all byproducts of incomplete combustion. Diesel particles have a carbon core wrapped in organic compounds and metals, while black carbon essentially acts as a delivery vehicle for these toxins. Together, they drive oxidative stress, inflammation, and cell damage (Kim et al., 2014; Janssen et al., 2011). The smaller particles are especially concerning because they pass the body’s usual defenses and get deep into the lungs (Wong et al., 2017; Ohlwein et al., 2019), and from there, along with the toxic compounds riding along with them, they can cross into the bloodstream and reach organs throughout the body (Bové et al., 2019).

Much of this damage stems from oxidative stress: too many reactive oxygen species (ROS) damage cells faster than the body can repair them (Li et al., 2018). Mitochondria are caught in the middle: they produce ROS and bear the brunt of the damage (Murphy et al., 2018; Picard & Shirihai, 2022). PM components interfere with mitochondrial respiration, disrupt mitochondrial membranes, and impair energy production (Kwon et al., 2020; Li et al., 2018). Black carbon’s reactive compounds and metals add to mitochondrial damage and energy disruption (Zong et al., 2024; Janssen et al., 2011). Together, these effects turn mitochondrial dysfunction into a central hub where all oxidative damage converges. Unlike nuclear DNA, mtDNA lacks histone protection and has limited repair capacity, making it especially exposed to oxidative damage (Gustafson et al., 2020). At first, cells might respond to oxidative stress by making more copies of their mtDNA (mtDNA-CN), a compensatory mechanism to keep energy production going (Castellani et al., 2020). But if the stress persists, it begins to damage the mtDNA itself, impairing replication and respiration and generating even more ROS, creating a self-perpetuating cycle (Picard & Shirihai, 2022; Gustafson et al., 2020). As a result, mtDNA-CN measured in blood has become a popular biomarker for mitochondrial health (Longchamps et al., 2020). It has been linked to aging, chronic disease, and poorer health outcomes generally (Ashar et al., 2017). Interestingly, PM exposure during pregnancy seems to have opposite effects depending on timing: a boost in mtDNA-CN early on, but a drop later in pregnancy.

These conflicting results likely reflect differences in PM exposure levels, particle size and composition, pollution sources, and the timing and duration of exposure during pregnancy. In addition, studies differ in populations, sample sizes, exposure measurement methods, biological samples used, mtDNA-CN quantification, and statistical methods, which could explain why findings don’t line up (Byun et al., 2013). Most research points to an inverse relationship between PM exposure and mtDNA-CN: more exposure, less mtDNA-CN, consistent with the idea that mitochondrial damage from oxidative stress accumulates over time (Hou et al., 2011). But some studies find the opposite, a positive association, which researchers think reflects an early adaptive response in which mitochondria ramp up production in response to oxidative stress (Lee & Wei, 2004; Castellani et al., 2020). For example, short- and medium-term exposure to black carbon has been linked to higher mtDNA-CN in older adults, with PM_₂.₅_ and PM_₁₀_ showing similar patterns, and physical activity appears to soften these effects somewhat (Zhong et al., 2015).

Despite this growing body of research, no one has synthesized it into a systematic review or meta-analysis, and no one has formally assessed the strength or reliability of the evidence using current evidence-grading frameworks. This study aims to fill that gap: we conducted a systematic review and meta-analysis to synthesize evidence on PM exposure and mtDNA-CN, evaluate the quality of individual studies, and assess the overall certainty of the evidence, providing a clearer picture of how PM exposure relates to mitochondrial dysfunction.

## 2. Methodology

This systematic review and meta-analysis followed the Preferred Reporting Items for Systematic Reviews and Meta-Analyses (PRISMA) guidelines and is registered with the PROSPERO database (No. CRD420261320957), keeping the review process transparent, reproducible, and well documented.

### 2.1. Literature search strategy

We conducted a systematic literature search in PubMed, Scopus, and Web of Science from each database’s inception through 20 February 2026 **(Supplementary Information)**. We developed the review objectives using the PECOS (Population, Exposure, Comparator, Outcome, and Study Design) framework to examine whether PM exposure is associated with changes in mtDNA-CN in both human and animal studies. We used a combination of MeSH terms and free-text keywords, connected with Boolean operators (AND, OR, and NOT), to find all relevant papers investigating ambient air pollution and mitochondrial biomarkers. The search terms covered ambient air pollution, PM (PM_₀.₁_, PM_₂.₅_, PM_₁₀_), DEP, PAHs, BC, mtDNA, mtDNA-CN, and mtDNA methylation. Full search strings are provided in the supplementary material **(Supplementary Material Tables S1-S8)**. We also manually screened reference lists of eligible articles. We screened review articles for citation tracking and excluded them from the final analysis.

### 2.2. Eligibility criteria and study selection

We included human epidemiological studies, preclinical animal studies, and relevant *in vitro* investigations from all geographic regions and age groups. This analysis examined the effects of exposure to ambient particulate matter (PM), including ultrafine particles (PM_₀.₁_), fine particles (PM_₂.₅_), coarse particles (PM_₁₀_), black carbon (BC), polycyclic aromatic hydrocarbons (PAHs), and diesel exhaust particles (DEPs) in both micro- and nanoscale forms. We included observational studies, animal experiments, and *in vitro* investigations examining the PM-mtDNA-CN relationship, provided they reported quantitative mtDNA-CN measurements from human samples, animal experiments, or *in vitro* work using human/rodent cell lines or primary cells exposed to realistic PM concentrations. We excluded reviews, editorials, commentaries, case reports, duplicate papers, and studies without sufficient quantitative data. We imported everything into Rayyan to manage references and remove duplicates **(Supplementary Table S1)**. Three reviewers (A.P., A.T., and A.K.S.) independently screened titles and abstracts, and relevant articles moved on to full-text review. The authors resolved disagreements through discussion, bringing in a fourth author (R.K.N.) when needed, and made final inclusion decisions by consensus.

### 2.3. Data extraction

The data covering study design, sample size, participant characteristics, biological matrix, exposure assessment, pollutant type and level, outcome measures, statistical models, and major findings were pulled by two authors (A.P., A.T.) using a standardized form **(Supplementary Table S2)**.

### 2.4. Assessment of risk of bias and study quality

A.P. and A.T. independently assessed methodological quality and risk of bias using the appropriate Joanna Briggs Institute (JBI) Critical Appraisal Tool for each study design. The appraisal considered design-specific domains relevant to methodological quality, including participant selection, exposure and outcome assessment, confounding, attrition or exclusion, and statistical analysis, where applicable. Because JBI tools differ by study design, the number and specific domains assessed varied across studies (**Table 2**). We also assessed evidence quality by examining consistency across included studies. We report detailed quality assessment scores and risk-of-bias judgments for each study **(Supplementary Table S6)**. We resolved discrepancies through discussion and brought in a third reviewer (A.K.S.) when we couldn’t reach consensus.

### 2.5. Data synthesis and meta-analysis

We determined exposure windows as in previous studies (Smith et al., 2023) and classified exposure periods as prenatal/parental, concurrent, or lagged. Across human, animal, and *in vitro* studies, we evaluated associations with PM_0.1_, PM_2.5_, PM_10_, PM_2.5-10_, and PM constituents such as BC, DEP, and PAHs. Most human birth cohort and animal studies have focused on PM exposure during pregnancy and its effect on offspring mtDNA copy number. We applied the following criteria to select eligible associations for the meta-analysis. For quantitative meta-analysis, we considered exposure-outcome associations reported in three or more independent studies investigating the same exposure-outcome relationship eligible for formal pooling. When fewer than three independent studies were available for a specific exposure subgroup, we retained the available estimates for descriptive synthesis and did not interpret them as formally pooled evidence (Hu et al., 2023; Thompson et al., 2022; Vrijheid et al., 2010). For consistency, where possible, we used associations from multi-pollutant models rather than single-pollutant models to adjust the exposure of interest for potential confounders and improve comparability across studies (Whaley et al., 2020). For each study, we converted β-coefficients or mean differences, where necessary, to a common effect metric and exposure increment for each pollutant. We calculated effect sizes to quantify the change in mtDNA- CN associated with the specified pollutant exposure. We combined all estimates within each exposure category and metric. Standard error estimation: Standard errors (*SE_i_*) were backcalculated from reported 95% confidence intervals (lower bound *L_i_*, upper bound *U_i_*) under the assumption of normality:

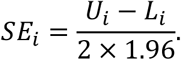

When unavailable, SEs were derived using Cochrane-recommended methods from *P*-values with effect estimates or from raw means and standard deviations.

Assessment of heterogeneity: Heterogeneity was evaluated using Cochran’s *Q*statistic and the *I*^2^index with fixed-effect inverse-variance weights (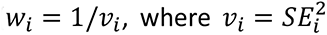). The fixed-effect pooled estimate was

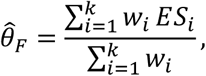

where *k* is the number of studies.

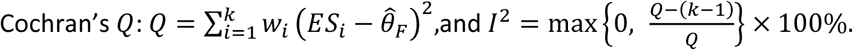

*I*^2^ > 50%indicated substantial heterogeneity and *I*^2^ > 75% Considerable heterogeneity. DerSimonian-Laird random-effects meta-analysis: We applied a DerSimonian-Laird random-effects model to account for anticipated heterogeneity arising from differences in exposures, methods, and populations (DerSimonian & Laird, 1986; Harrer et al., 2021). Between-study variance *τ*^2^was estimated via the scaling factor.

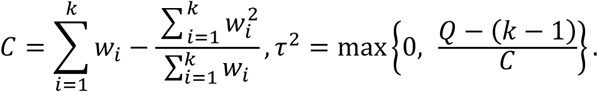

Random-effects weights were

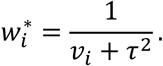

The pooled estimate was

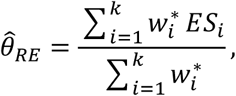

with standard error

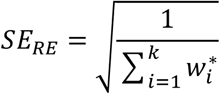

and 95% confidence limits θ̂*_RE_* ± 1.96 × *SE_RE_*. A pooled effect was statistically significant (*α* = 0.05) if the CI excluded zero.

Statistical significance testing: Significance of the pooled effect was assessed with a *z*-test:

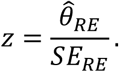

A two-tailed *p*-value was obtained from the standard normal cumulative distribution function; significance was defined as *p* < 0.05.

We assessed potential publication bias by visually inspecting funnel plots. We conducted subgroup analyses by species (human versus animal models) and PM exposure type (PM₂.₅, PM₁₀, and mixed PM exposure) to explore potential sources of heterogeneity and assess whether these factors modified the association between PM exposure and mtDNA-CN. In addition to mtDNA-CN, related mitochondrial and oxidative-stress endpoints reported across the included studies, mtDNA methylation, mitochondrial biogenesis markers, inflammation, and ROS were incorporated as subgroup dimensions to examine whether PM exposure effects differed across these mechanistically linked outcomes.

## 3. Results

### 3.1. Study selection and characteristics

We searched three major databases, PubMed, Scopus, and Web of Science, and found 1,224 records to start (320 from PubMed, 624 from Scopus, and 280 from Web of Science) (**Figure 1**). After removing 341 duplicates, we screened 883 unique records by title and abstract, narrowing to 143 potentially eligible articles for full-text review. We assessed each study’s relevance, design, methodology, and eligibility. This left 48 studies meeting our basic criteria; of those, 24 had enough quantitative data to include in the systematic review and meta-analysis **(Supplementary Table S3)**.

**Figure 1:**
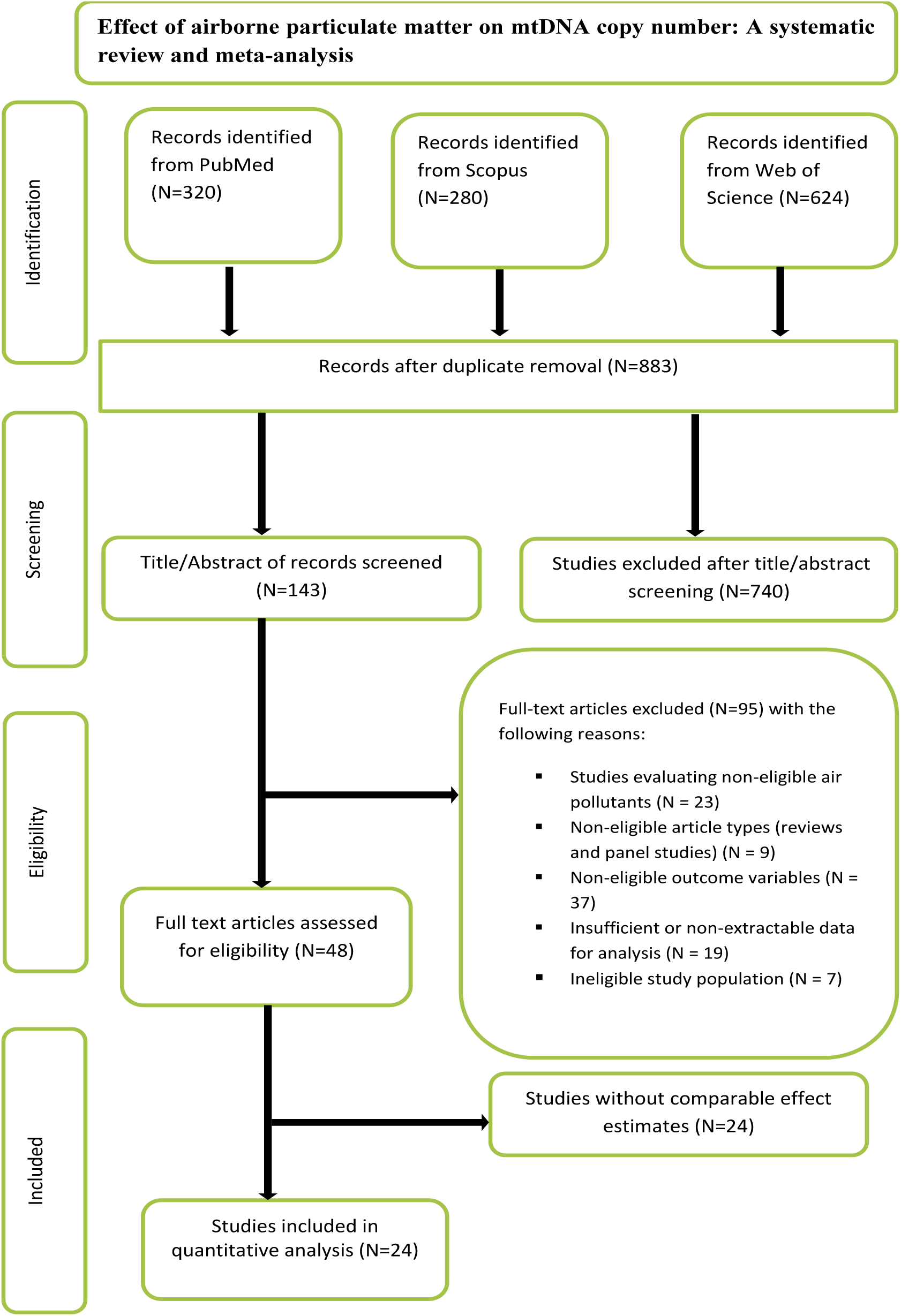
PRISMA flow diagram of study selection.

The 24 studies included in the quantitative synthesis examined a range of ambient air pollution exposures and their associations with mtDNA copy number (mtDNA-CN). Of these, three studies on general particulate matter (PM) exposure (Brunst et al., 2017; Van der Stukken et al., 2022; Hou et al., 2010), whereas six focused specifically on fine particulate matter (PM_₂.₅_) (Hou et al., 2013; Rosa et al., 2016; Janssen et al., 2015; Mandakh et al., 2021; Kaali et al., 2018; Peng et al., 2017). Four zeroed in on PM₁₀ (Janssen et al., 2012; Clemente et al., 2017; Clemente et al., 2016; Gaikwad et al., 2020), and another four looked at both PM_₂.₅_ and PM_₁₀_ together (Zhang et al., 2020; Wang et al., 2020; Hu et al., 2020; Grevendonk et al., 2016). One study included PM_₀.₁_ alongside PM₂.₅ and PM_₁₀_ (Li et al., 2021). One study reported black carbon alone (Bai et al., 2018), and three others reported it alongside PM_₂.₅_ (Hautekiet et al., 2021; Hautekiet et al., 2025; Vos et al., 2020). Two studies looked at PAH exposure (Pieters et al., 2013; Carugno et al., 2011). We ran all 24 studies through the JBI critical appraisal tool for quality, and every single one scored at least 50%, so all 24 made it into our final evidence base. Of these, 12 reported mtDNA-CN as a percentage change and 12 as an absolute value change, and we included both types in the quantitative meta-analysis. We excluded another 24 studies from the meta-analysis because they reported only qualitative, directional findings and lacked sufficient quantitative data to pool. This section summarizes the key features of the included studies, which used cross-sectional, cohort, experimental, and observational designs. The studies were published between 2010 and 2025, with study or exposure periods extending back to 1989. Peripheral blood was by far the most common sample type used to measure mtDNA-CN, followed by cord blood, placental tissue, sperm, buccal cells, neuronal cells, and epithelial cells. Geographically, Belgium contributed the most studies (n = 10), followed by China (n = 4), the United States (n = 3), and Italy (n = 2), with one study each from Mexico, India, Spain, Sweden, and Ghana (**Table 1**).

**Table 1.**
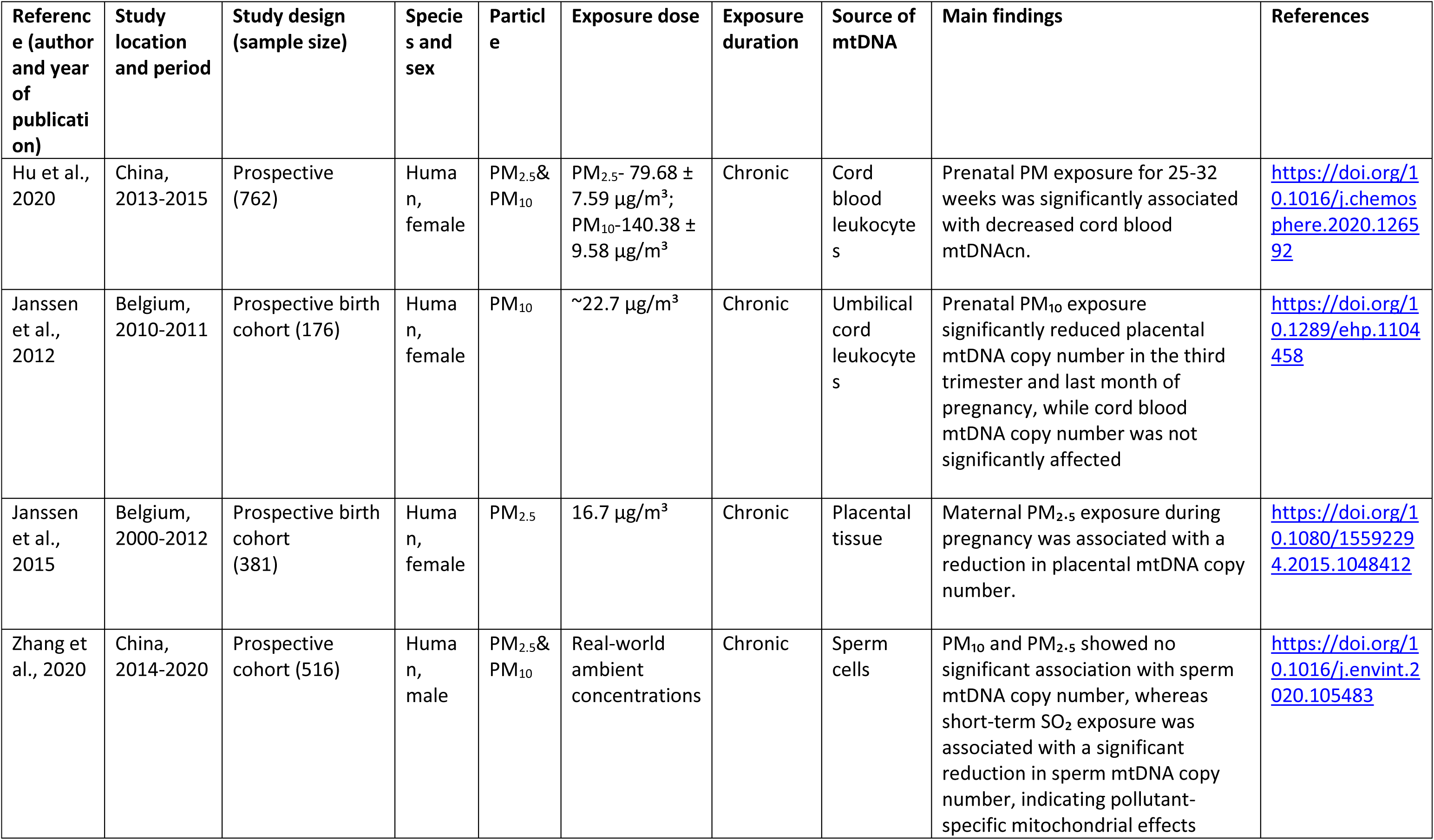
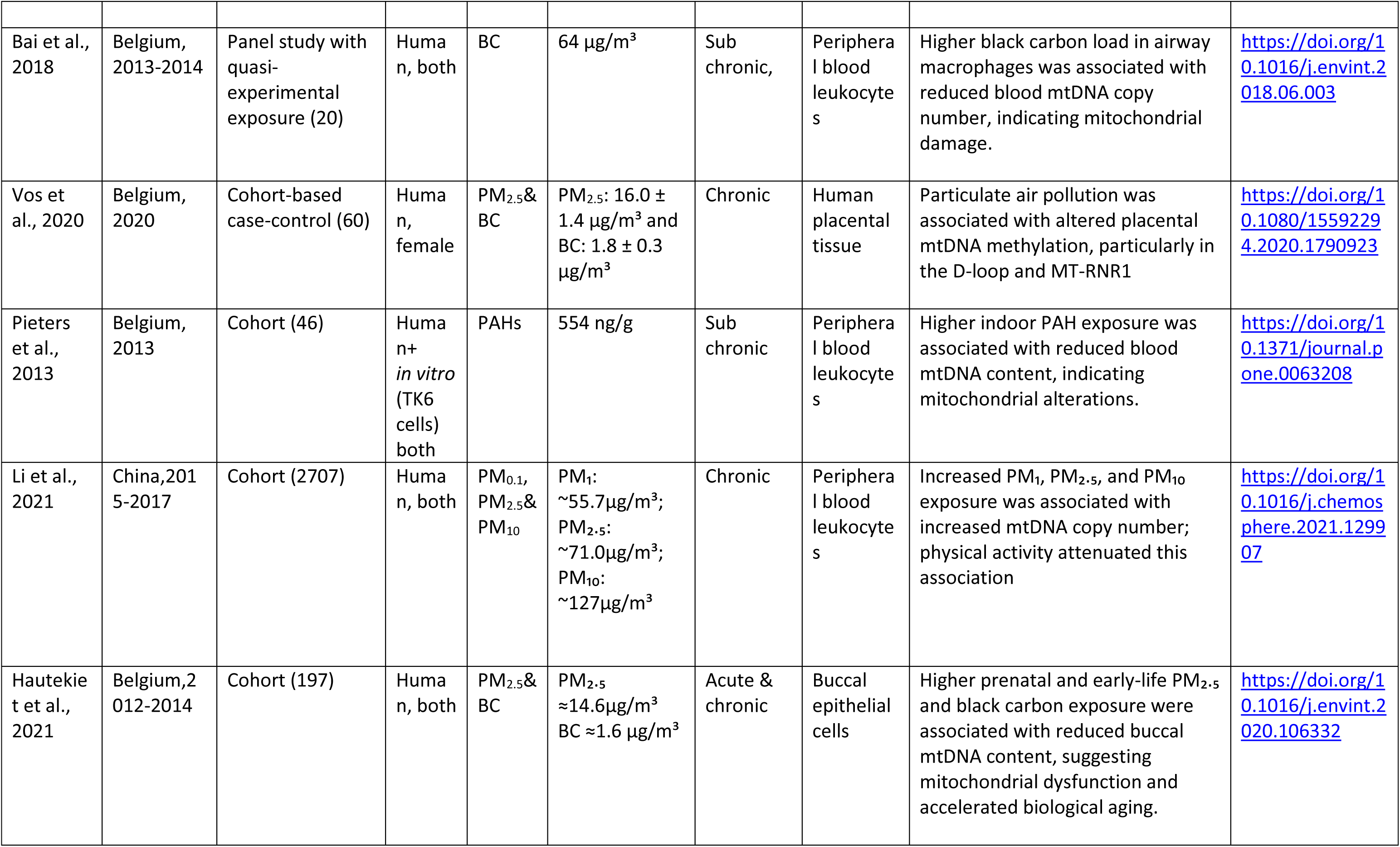
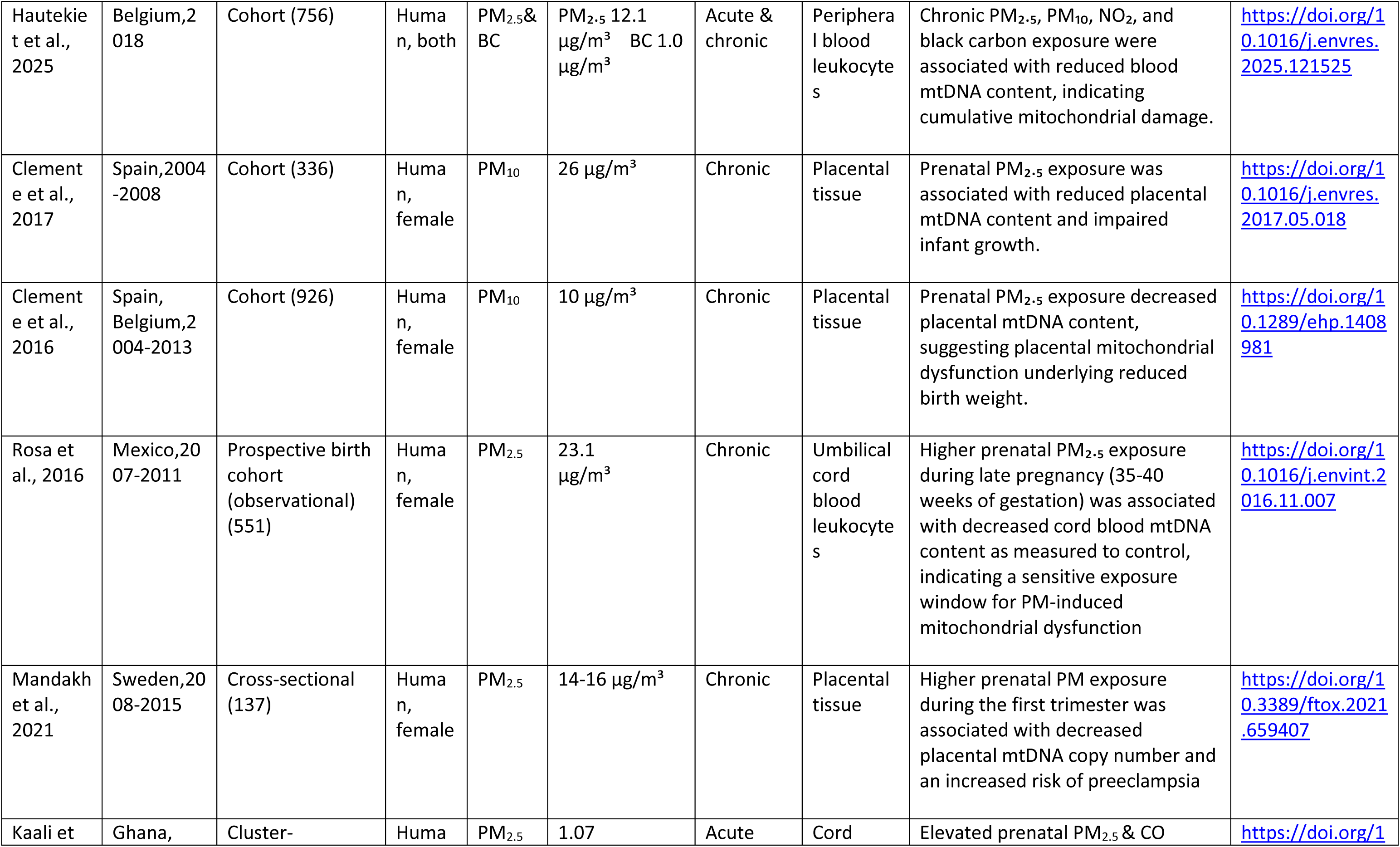
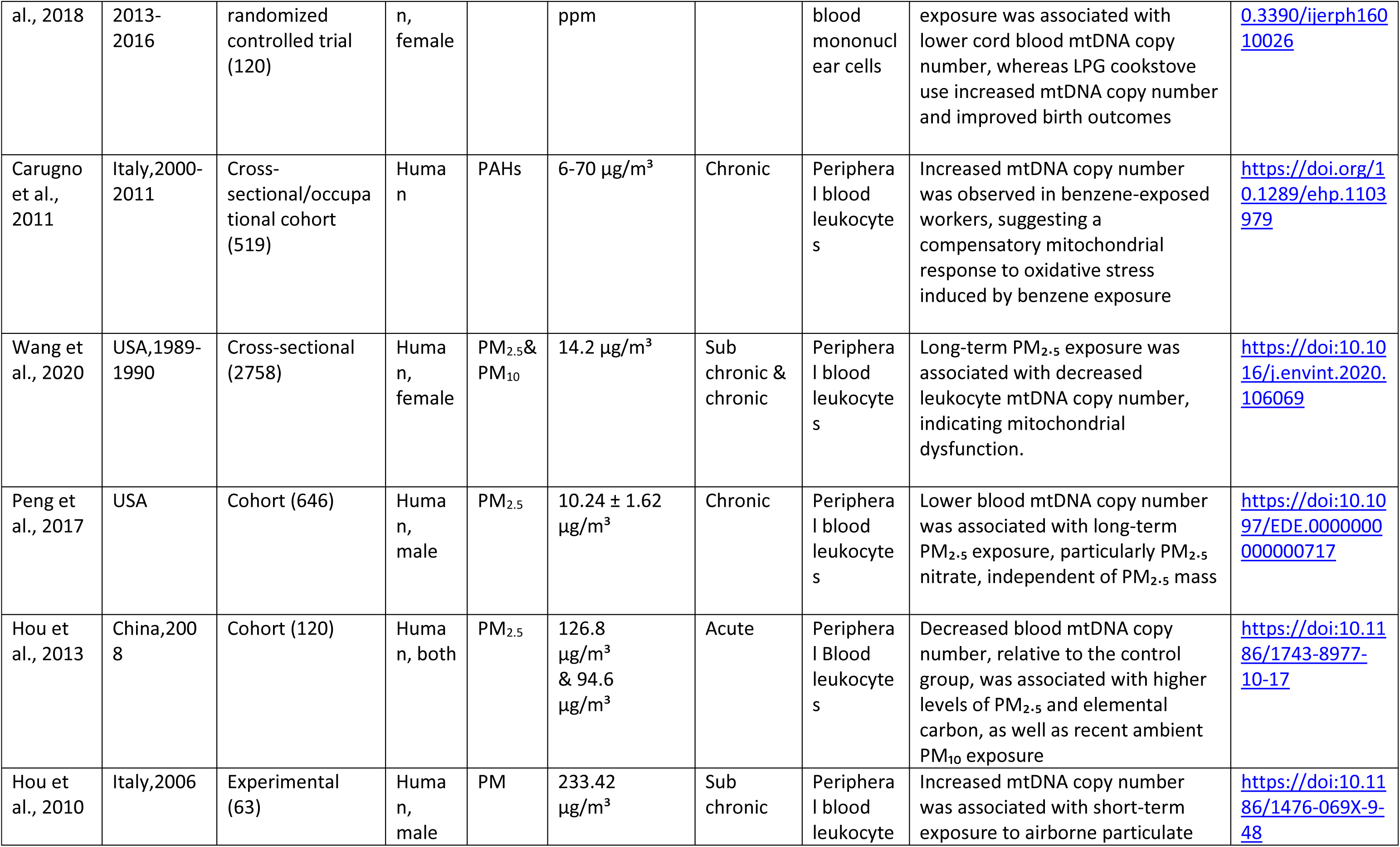
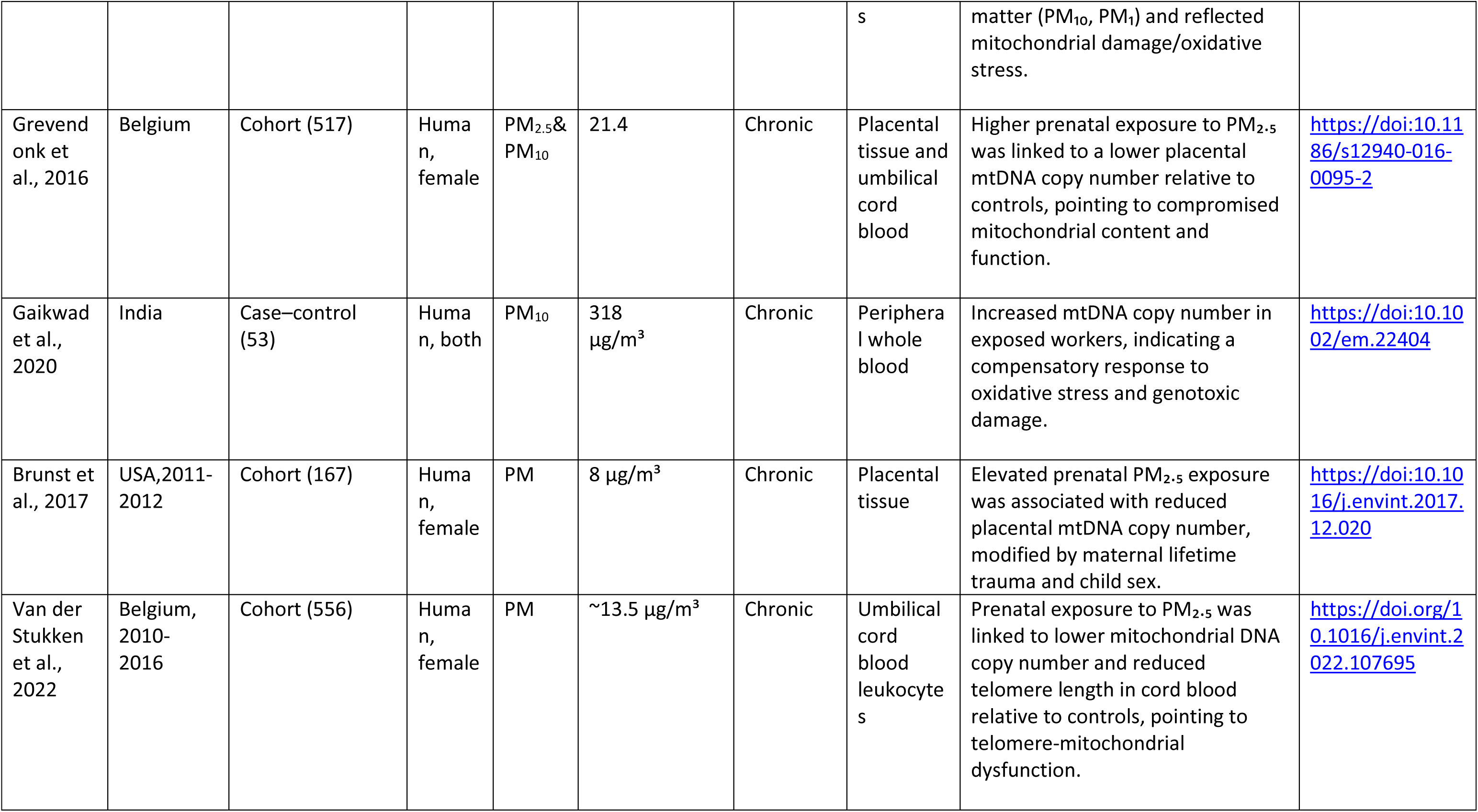
Characteristics and main findings of studies evaluating the effects of particulate matter (PM) exposure on mitochondrial DNA copy number (mtDNA-CN).

**Table 2:**
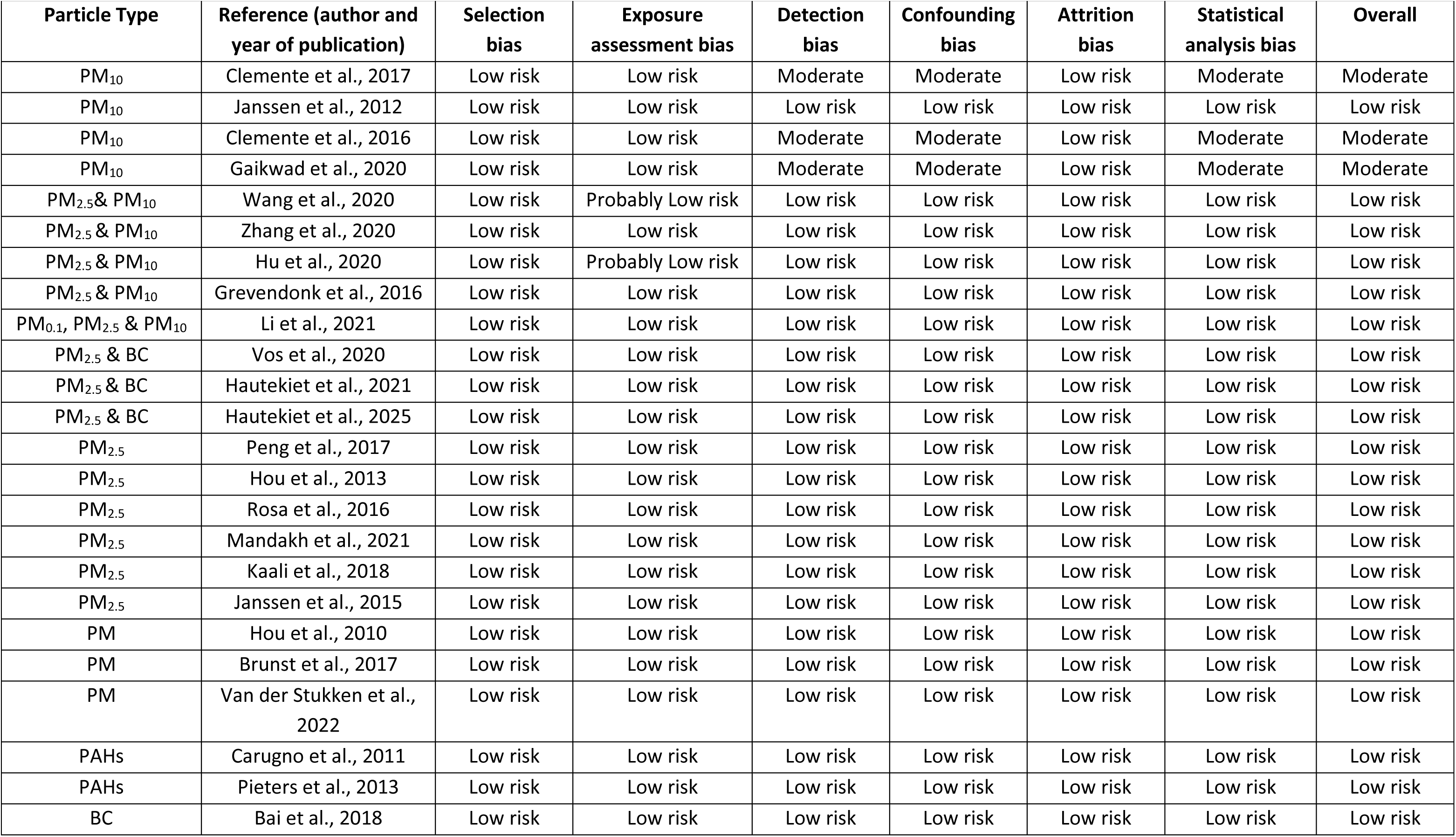
Assessment of Risk of Bias using Joanna Briggs Institute (JBI) Critical Appraisal Tools for bias approach.

### 3.2. Risk of bias assessment

Overall, the 24 studies performed well in terms of risk of bias: 87.5% (n = 21) were classified as low risk, 12.5% (n = 3) as moderate, and none as high risk (**Table 2**). Selection bias was the primary issue in the moderate-risk studies, usually because they recruited certain subgroups rather than generally representative samples. The use of approved exposure methodologies kept detection bias low. Standardized lab procedures and consistent reporting supported the generally low levels of outcome evaluation and reporting bias **(Supplementary Table S7)**.

### 3.3. Meta-analysis of the association between ambient PM and mtDNA-CN

Twelve studies reported percentage changes in mtDNA-CN and 12 reported absolute changes. We synthesized these estimates separately, with formal subgroup pooling restricted to exposure categories supported by at least three independent studies; we retained single-study and two-study subgroup findings for descriptive synthesis. Twelve studies reported percentage changes in mtDNA- CN and 12 reported absolute changes, which were synthesized separately. The percentage-change analysis yielded an overall reduction in mtDNA-CN (ES = −4.90, 95% CI: −7.97 to −1.82, p = 0.002), whereas the absolute-change analysis yielded a small overall increase (ES = 0.55, 95% CI: 0.05 to 1.04, p = 0.030). Both analyses showed considerable between-study heterogeneity (I² > 90%), so interpret the pooled estimates cautiously.

#### 3.3.1. Subgroup analysis by pollutant type: absolute value change in mtDNA-CN

Two studies investigated PAH exposure; however, only one contributed to the absolute-change analysis. We therefore interpreted this finding descriptively rather than as a formally pooled association. Two studies contributed to the general PM subgroup and reported divergent findings; we interpreted this subgroup descriptively as well. Similarly, two studies contributed to the combined PM_₂.₅_/PM_₁₀_ subgroup and reported divergent findings; we did not consider this subgroup a formal pooled association **(Supplementary Table S4**) (**Figure 2**).

**Figure 2.**
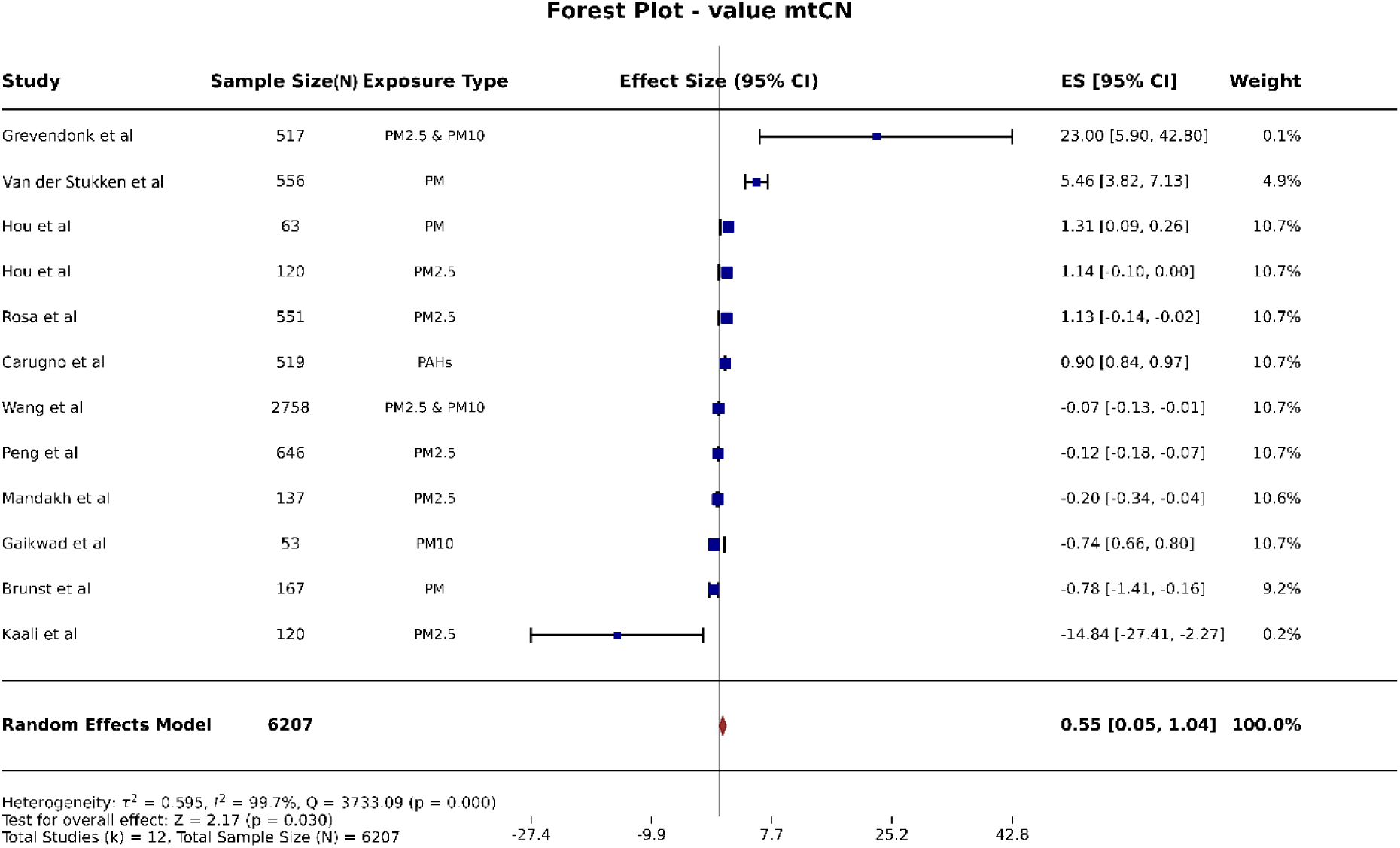
Forest plot showing the association between air pollutant exposure and absolute change in mtDNA copy number (mtDNA-CN). Squares represent individual study effect sizes with 95% confidence intervals (sized by study weight). The diamond shows the random-effects pooled estimate (ES = 0.55, 95% CI: 0.05–1.04; p = 0.030; N = 6207; k = 12). Heterogeneity was high (I² = 99.7%).

#### 3.3.2. Subgroup analysis by pollutant type: percentage change in mtDNA-CN

Exposure to PM_₂.₅_ alone was linked to a clear drop in mtDNA-CN (ES = −15.60), and the same was true for combined PM_₂.₅_ and PM_₁₀_ exposure (ES = −8.69, p < 0.001). Only one study contributed to the BC subgroup; therefore, we interpreted this finding descriptively rather than as a formally pooled association. By contrast, PM_₁₀_ alone (ES = −3.48, p = 0.497) and combined PM_₂.₅_ + BC exposure (ES = −3.01, p = 0.139) showed no significant association, possibly because of high heterogeneity in those subgroups. Only one study examined the combined PM_₀.₁_, PM_₂.₅_, and PM_₁₀_ exposure subgroup; therefore, we treated this finding as a descriptive observation rather than a formally pooled association **(Supplementary Table S5**) (**Figure 3**).

**Figure 3.**
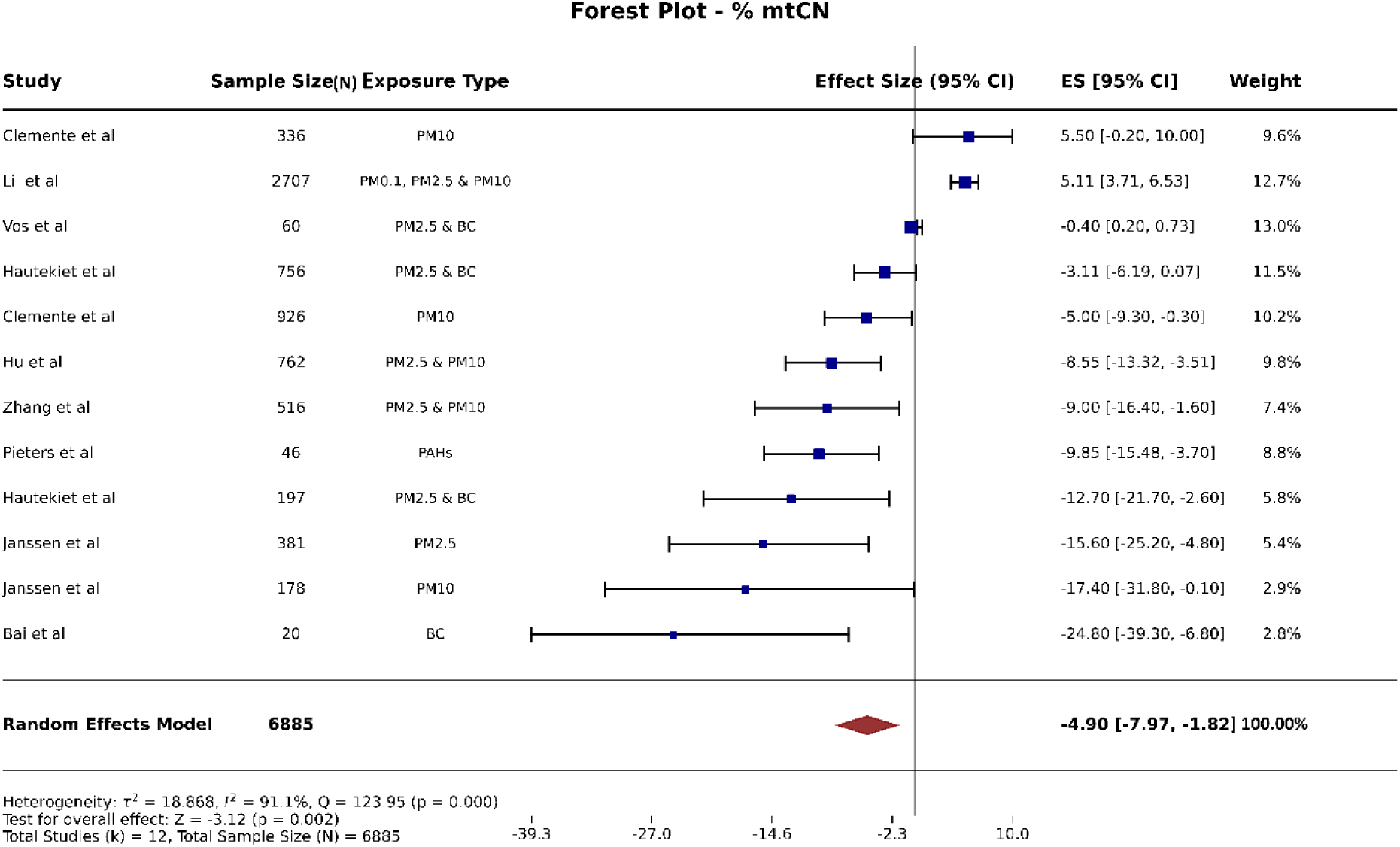
Forest plot depicting the relationship between PM exposure and % change in mtDNA-CN. Squares represent individual study effect sizes with 95% confidence intervals (sized by study weight). The diamond shows the random-effects pooled estimate (ES = −4.90, 95% CI: −7.97 to −1.82; p = 0.002; N = 6885; k = 12). Heterogeneity was high (I² = 91.1%).

#### 3.3.3. Subgroup analysis: mtDNA methylation

Five of the 48 eligible studies examined mtDNA methylation in relation to particulate matter exposure (**Table 3**). We included these studies in the mechanistic synthesis, regardless of whether they contributed quantitative mtDNA-CN data to the meta-analysis. Four studies found hypermethylation in key mitochondrial regulatory regions, the D-loop, MT-RNR1, and MT-TF, alongside lower mtDNA-CN after PM_₂.₅_ or BC exposure. Bhargava et al. (2020) were the odd one out, finding global mtDNA hypomethylation tied to PAH exposure, even though mtDNA-CN was still reduced. Taken together, these results suggest that hypermethylation of mitochondrial regulatory regions is the more typical epigenetic response to particulate air pollution, and it usually goes hand in hand with reduced mtDNA-CN, a sign of impaired mitochondrial function.

**Table 3.** Overview of studies examining mitochondrial DNA methylation in relation to particulate matter (PM) exposure.

| Reference (author and year of publication) | Study location and period | Study design | Species & sample size | Particle | Exposure duration | mtDNA methylation (yes/no) | Regions analysed | Increase/decrease | Main findings | References |
| --- | --- | --- | --- | --- | --- | --- | --- | --- | --- | --- |
| Janssen et al., 2015 | Belgium, 2000-2012 | Prospective birth cohort (observational) | Human (381) | PM 2.5 | Chronic | Yes | D-loop, and MT-RNR1(12S rRNA) | Increase | Higher prenatal PM<sub>2.5</sub> exposure was associated with mtDNA hypermethylation (D-loop and MT-RNR1) and decreased placental mtDNA content. | https://doi.org/10.1080/15592294.2015.1048412 |
| Mishra et al., 2022 | India, 2017-2018 | Cross-sectional observational | Human (120) | PM 2.5 | Chronic | Yes | D-loop1, D-loop2, 12S+TF, CYTB, 16S | Increase | PM exposure induced hypermethylation of mitochondrial regulatory regions, accompanied by oxidative stress, mitochondrial dysfunction, and impaired mitochondrial biogenesis. | https://doi.org/10.1016/j.apr.2022.101399 |
| Byun et al., 2013 | Italy and China | Observational | Human (120) | PM | Chronic | Yes | D-loop, MT-TF, MT-RNR1 | Increase | Metal-rich PM<sub>1</sub> exposure induced locus-specific mtDNA hypermethylation (MT-TF and MT-RNR1) without affecting the D-loop | https://doi.org/10.1186/1743-8977-10-18 |
| Vos et al., 2020 | Belgium, 2020 | Cohort-based case-control | Human (60) | PM<sub>2.5</sub> & BC | Chronic | Yes | D-loop and MT-RNR1 | Increase | Prenatal environmental stressors increased placental D-loop and MT-RNR1 methylation, highlighting mtDNA methylation as an early biomarker of prenatal exposure | https://doi.org/10.1080/15592294.2020.1790923 |
| Bhargava et al., 2020 | India, 2020 | Experimental | Human cell line | PAH | Sub chronic | Yes | Global mtDNA | Decrease | BaP induced global mtDNA hypomethylation, whereas anthracene caused no significant mtDNA methylation changes. | https://doi.org/10.1177/1091581820932875 |

#### 3.3.4. Subgroup analysis: mitochondrial biogenesis, inflammation, and dynamics

Of the 48 eligible studies, six unique studies investigated mechanistic pathways; however, Mishra et al. (2022) contributed findings across two mechanistic domains, mitochondrial biogenesis and mitochondrial dynamics, resulting in seven study-domain entries in **Table 4**. PM₂.₅ or PAH exposure was consistently linked to lower PGC-1α and TFAM expression alongside reduced mtDNA-CN, suggesting impaired mitochondrial biogenesis. One study bucked the trend, showing increased TFAM expression, which might reflect a compensatory response rather than a consistent pattern. Other findings pointed to ROS-driven activation of NF-κB signaling, higher levels of pro-inflammatory cytokines (TNF-α, IL-1β, and IL-6), and disrupted mitochondrial dynamics, shown by increased DRP1 alongside decreased MFN2 and OPA1. Altogether, these findings suggest that PM-related drops in mtDNA-CN are associated with impaired mitochondrial biogenesis, increased inflammation, and a disrupted balance between mitochondrial fission and fusion.

**Table 4.** Studies evaluating mitochondrial biogenesis, mitochondrial dynamics, and inflammatory signaling in relation to particulate matter (PM) exposure.

| Reference (author and year of publication) | Study location and period | Study design | Species & sample size | Particle | Exposure duration | Mitochondrial biogenesis PGC-1α/TFAM (yes/no) | Increase/decrease | Main findings | References |
| --- | --- | --- | --- | --- | --- | --- | --- | --- | --- |
| Sivakumar et al.,2024 | India, 2023-2024 | Experimental | Rat (24) | PM<sub>2.5</sub> | Sub chronic | Assessed both TFAM & PGC-1α | Decrease | PM<sub>2.5</sub> exposure downregulated PGC-1α and TFAM, leading to decreased mtDNA copy number and impaired mitochondrial biogenesis in the hearts of uremic cardiomyopathic rats. | https://doi.org/10.1016/j.envpo.2024.124113 |
| Chang-Chien et al.,2024 | Taiwan,2024 | Experimental | Human (BEAS-2B) | PM 2.5 and PM 10 | Acute | Assessed both TFAM & PGC-1α | Decrease | PM exposure decreased PGC-1α, TFAM, and mtDNA copy number, an effect reversed by vitamin D. | https://doi.org/10.1186/s12931-024-02951-7 |
| Bhargava et al.,2020 | India,2020 | Experimental | Human | PAHs | Acute & sub chronic | Assessed both TFAM & PGC-1α | Decrease | Carcinogenic PAH exposure decreased PGC-1α, TFAM, and mtDNA copy number, impairing mitochondrial biogenesis via oxidative stress-induced epigenetic alterations. | https://doi.org/10.1177/1091581820932875 |
| Mishra et al.,2022<br>Mishra et al., 2022 | India,2017-2018 | Cross-sectional observational | Human (120) | PM 2.5 | Chronic | Assessed TFAM only | Increase | Airborne PM exposure increased TFAM expression, altered mtDNA copy number, and induced mtDNA hypermethylation | https://doi.org/10.1016/j.apr.2022.101399 |
|  | India | Cross-sectional observational | Human | PM 2.5 | Chronic | Yes analysed | DRP increased, and MFN and OPA1 decreased | Airborne PM exposure increased DRP1 and decreased MFN2 and OPA1, disrupting mitochondrial dynamics and promoting mitochondrial dysfunction through integrated mitoeipigenetic | https://doi.org/10.1016/j.apr.2022.101399 |

|  |  |  |  |  |  |  |  | signaling. |  |
| --- | --- | --- | --- | --- | --- | --- | --- | --- | --- |
| Lin et al.,2022 | Taiwan, 2021 | Experimental | Human | PM 2.5 | Acute | Assessed (NF-κB, TNF-α, IL-1β). | increase | PM<sub>2.5</sub> exposure induced oxidative stress by increasing ROS levels and activating NF-κB signaling, resulting in elevated TNF-α and IL-1β expression and enhanced inflammation | https://doi.org/10.1016/j.neuro.2021.10.009 |
| Bai et al., 2018 | Belgium, 2013-2014 | Panel study with quasi-experimental exposure contrast | Human (20) | BC | Sub chronic | Assessed (IL-6). | increase | mtDNA content showed an inverse association with inflammatory markers, including hs-CRP, hs-IL-6 | https://doi.org/10.1016/j.envint.2018.06.003 |

### 3.4. Confidence in the body of evidence and level of evidence

Overall, we rated the certainty of the evidence as low, reflecting the predominantly observational nature of the included studies, considerable between-study heterogeneity, and variability in exposure assessment and mtDNA-CN measurement methods. PM₂.₅ showed a significant inverse association (ES = −15.60), but certainty was low given the high heterogeneity (I² > 90%; six studies). PM₁₀ came out non-significant (ES = −3.48, p = 0.497), also with low certainty.

## 4. Discussion

This review synthesized evidence from 24 quantitative studies looking at how ambient PM and related pollutants relate to mtDNA-CN, along with mechanistic evidence on mitochondrial epigenetic changes, bioenergetic dysfunction, and inflammation. Overall, pollutant exposure was tied to a significant drop in percentage-based mtDNA-CN, but a modest, significant increase when studies reported absolute values instead. Both pooled estimates showed substantial heterogeneity (I² > 90%), which tracks with how much pollutant types, exposure metrics, study populations, and methods varied. Breaking things down by subgroup, finer particle fractions and specific components - especially PM₂.₅, black carbon, and PAHs showed up more consistently linked to lower % mtDNA-CN. The absolute-value results were shakier: PAH exposure alone looked linked to an increase, but that’s based on just two studies, so it’s worth taking with a grain of salt. Broader PM categories, meanwhile, gave non-significant and inconsistent results. Combined PM₂.₅ and BC exposure was tied to a real drop in mtDNA-CN, though BC alone wasn’t. And when we pooled everything together regardless of how it was reported, the overall association disappeared, which just reflects how much heterogeneity there is, not that there’s no real biological effect. These results imply that the choice of mtDNA-CN measurement metric can affect the observed relationships, and that finer PM and specific chemical elements have more detrimental impacts on mitochondrial health than broader PM categories.

The drop in mtDNA-CN after PM exposure makes biological sense and fits with what we already know about how particulates cause harm (Cheng et al., 2024). Fine and ultrafine particles can cross the alveolar barrier and get into the bloodstream, carrying toxic components like transition metals, black carbon, diesel exhaust particles, and PAHs - all of which drive excessive production of ROS (Ding et al., 2025). Persistent oxidative stress damages the respiratory chain, impairs oxidative phosphorylation, disrupts mitochondrial replication, and eventually reduces mtDNA-CN, reflecting the gradual accumulation of mitochondrial damage (Yin & Guo, 2025). Most studies found this inverse pattern; a few saw mtDNA-CN increase after particulate matter exposure (Qiao et al., 2024), suggesting mitochondria don’t always respond the same way; it likely depends on the specifics of the exposure and the biological context. That inconsistency likely reflects differences in exposure duration, pollutant composition, exposure intensity, sample type, and study population. Early on, or with lower-level oxidative stress, cells might respond by ramping up mitochondrial biogenesis and switching on regulators like PGC-1α, NRF1, and TFAM, leading to a temporary bump in mtDNA-CN (Morris et al., 2024). But if exposure continues or intensifies, this adaptive response may be overwhelmed, leading to impaired biogenesis, reduced ATP production, and eventual drops in mtDNA-CN (Gustafsson et al., 2016).

Beyond mtDNA-CN itself, this review also turned up interesting evidence on mitochondrial epigenetics. Of the five studies that examined mtDNA methylation, four found hypermethylation after PM exposure, and only one found hypomethylation. The biological significance of mtDNA methylation is not yet fully understood; growing evidence suggests that methylation of the mitochondrial D-loop can inhibit mtDNA replication and transcription (Stoccoro et al., 2021), which in turn reduces mitochondrial gene expression and impairs oxidative phosphorylation (Yin et al., 2025). Environmental pollutants have been shown before to alter mitochondrial methylation by triggering DNA methyltransferases through oxidative stress, suggesting PM might cause lasting epigenetic changes in mitochondria (Byun et al., 2013). All this supports the idea that changes in mtDNA methylation might be an upstream driver behind the mtDNA-CN reductions we’re seeing across these studies.

The mechanistic evidence also shows PM throws off mitochondrial energy production more broadly. Most studies looking at mitochondrial biogenesis found that lower PGC-1α and TFAM are two key regulators that coordinate mitochondrial replication, transcription, and respiration- which lines up with impaired biogenesis, reduced ATP, and a harder time keeping cellular energy balanced (Chen et al., 2022). Interestingly, one study found the opposite higher expression of these regulators which reinforces the idea that mitochondrial responses depend a lot on how long and how severe the exposure is. Changes in the proteins that control mitochondrial fission and fusion (higher DRP1, lower MFN2 and OPA1) also point to a disrupted balance there (Xu et al., 2025). Keeping fission and fusion in balance matters for quality control, mitophagy, and overall energy production (Chen et al., 2023).

Inflammation is another link between PM exposure and mitochondrial dysfunction (Lin et al., 2022). Mechanistic studies consistently show activation of the NF-κB/NLRP3 inflammasome pathway after particulate exposure. Excess mitochondrial ROS triggers NF-κB activation, which increases production of inflammatory cytokines such as IL-1β, IL-6, and TNF-α (Mittal et al., 2013). At the same time, damaged mitochondria release oxidized mtDNA and other danger signals that activate the NLRP3 inflammasome (Qiu et al., 2022). These inflammatory responses then feed back into more oxidative stress and mitochondrial damage, a self-reinforcing cycle that drives chronic inflammation and tissue damage (Mittal et al., 2013). This meta-analysis shows substantial heterogeneity (I² > 90%) across the pooled results. Differences in pollutant makeup, exposure windows, biological samples (blood, placenta, cord blood, sperm, buccal cells), lab methods, and study populations almost certainly drove much of this and probably explain why the size and sometimes even the direction of the reported associations varied so much. By reporting format, we found significant pooled effects for studies reporting percentage changes, but not for those reporting absolute values. That should be read with some caution, but it hints that how studies analyze and normalize their data might genuinely matter for the numbers you end up with. Standardizing how mtDNA-CN is measured, how exposure is assessed, and how results are reported would go a long way toward making future studies more comparable and reducing this kind of heterogeneity.

This review has some real strengths: a dual-metric approach to the meta-analysis, a thorough quality check using the JBI tool (every study scored at least 50%), and integration of mechanistic evidence on methylation, biogenesis, inflammation, and mitochondrial dynamics. That said, there are limitations too: we couldn’t pool the qualitative studies, publication bias may have occurred, exposure-response relationships weren’t always fully reported, and we didn’t have individual-participant data that would’ve let us adjust for covariates more precisely. The evidence suggests that oxidative stress, epigenetic modifications, and reduced biogenesis are the major effects of ambient PM and associated contaminants that damage mitochondrial genome material. Further progress will require large, well-designed prospective studies with standardized exposure assessment and mtDNA-CN measurement methods, together with integrated mitoepigenetic and functional analyses.

## 5. Conclusion

The 24 studies included in this systematic review provided consistent evidence that exposure to ambient particulate matter is associated with decreased mtDNA-CN for PM₂.₅, BC, and PAHs. All included studies found a significant association between air pollution exposure and reduced mtDNA- CN, expressed as a percent change. Several plausible biological mechanisms may explain the associations observed, including oxidative stress, mtDNA hypermethylation, mitochondrial biogenesis defects, altered mitochondrial dynamics, and inflammatory pathways. Future research should focus on large prospective cohort studies and standardizing exposure assessment and mtDNA-CN measurement methods to provide more robust evidence on causal relationships and establish mtDNA-CN as a biomarker of air pollution exposure.

## Ethics

The study received approval from the Institutional Ethics Committee (IEC) of ICMR-NIREH under reference number INTR-IM-2024-00063 (PKM).

## Data availability

Data will be made available on request.

## Supplementary Information

Additional supporting information for this manuscript is available as **Supplementary Tables S1-S8**. The complete supplementary file can be accessed using the Supplementary Information hyperlink associated with this manuscript on the medRxiv submission page.

## Data Availability

All data generated or analyzed during this systematic review and meta-analysis are available from the corresponding author upon reasonable request. The study protocol, data extraction forms, extracted data, and statistical analysis files are available upon request.

https://www.crd.york.ac.uk/PROSPERO/view/CRD420261320957

## Acknowledgement

The first author (AP) gratefully acknowledges the Council of Scientific & Industrial Research (CSIR) for providing financial assistance through a Junior Research Fellowship (ID: 35112152). The authors thank Dr. Debabrata Dash for critical inputs.

## References

1. Ashar, F. N., Zhang, Y., Longchamps, R. J., Lane, J., Moes, A., Grove, M. L., Mychaleckyj, J. C., Taylor, K. D., Coresh, J., Rotter, J. I., Boerwinkle, E., Pankratz, N., Guallar, E., & Arking, D. E. (2017). Association of mitochondrial DNA copy number with cardiovascular disease. JAMA Cardiology, 2(11), 1247. 10.1001/jamacardio.2017.3683

2. Bai, Y., Casas, L., Scheers, H., Janssen, B. G., Nemery, B., & Nawrot, T. S. (2018). Mitochondrial DNA content in blood and carbon load in airway macrophages. A panel study in elderly subjects. Environment International, 119, 47–53. 10.1016/j.envint.2018.06.003

3. Bhargava, A., Kumari, R., Khare, S., Shandilya, R., Gupta, P. K., Tiwari, R., Rahman, A., Chaudhury, K., Goryacheva, I. Y., & Mishra, P. K. (2020). Mapping the mitochondrial regulation of epigenetic modifications in association with carcinogenic and noncarcinogenic polycyclic aromatic hydrocarbon exposure. International Journal of Toxicology, 39(5), 465–476. 10.1177/1091581820932875

4. Bové, H., Bongaerts, E., Slenders, E., Bijnens, E. M., Saenen, N. D., Gyselaers, W., Van Eyken, P., Plusquin, M., Roeffaers, M. B. J., Ameloot, M., & Nawrot, T. S. (2019). Ambient black carbon particles reach the fetal side of human placenta. Nature Communications, 10(1), 3866. 10.1038/s41467-019-11654-3

5. Brunst, K. J., Sanchez-Guerra, M., Chiu, Y. M., Wilson, A., Coull, B. A., Kloog, I., Schwartz, J., Brennan, K. J., Enlow, M. B., Wright, R. O., Baccarelli, A. A., & Wright, R. J. (2017). Prenatal particulate matter exposure and mitochondrial dysfunction at the maternal-fetal interface: Effect modification by maternal lifetime trauma and child sex. Environment International, 112, 49–58. 10.1016/j.envint.2017.12.020

6. Byun, H., Panni, T., Motta, V., Hou, L., Nordio, F., Apostoli, P., Bertazzi, P., & Baccarelli, A. A. (2013). Effects of airborne pollutants on mitochondrial DNA Methylation. Particle and Fibre Toxicology, 10(1), 18. 10.1186/1743-8977-10-18

7. Carugno, M., Pesatori, A. C., Dioni, L., Hoxha, M., Bollati, V., Albetti, B., Byun, H., Bonzini, M., Fustinoni, S., Cocco, P., Satta, G., Zucca, M., Merlo, D. F., Cipolla, M., Bertazzi, P. A., & Baccarelli, A. (2011). Increased Mitochondrial DNA Copy Number in Occupations Associated with Low-Dose Benzene Exposure. Environmental Health Perspectives, 120(2), 210–215. 10.1289/ehp.1103979

8. Castellani, C. A., Longchamps, R. J., Sun, J., Guallar, E., & Arking, D. E. (2020). Thinking outside the nucleus: Mitochondrial DNA copy number in health and disease. Mitochondrion, 53, 214–223. 10.1016/j.mito.2020.06.004

9. Chang-Chien, J., Huang, J., Tsai, H., Wang, S., Kuo, M., & Yao, T. (2024). Vitamin D ameliorates particulate matter induced mitochondrial damages and calcium dyshomeostasis in BEAS-2B human bronchial epithelial cells. Respiratory Research, 25(1), 321. 10.1186/s12931-024-02951-7

10. Chen, L., Qin, Y., Liu, B., Gao, M., Li, A., Li, X., & Gong, G. (2022). PGC-1Α-Mediated Mitochondrial Quality control: Molecular mechanisms and implications for heart failure. Frontiers in Cell and Developmental Biology, 10, 871357. 10.3389/fcell.2022.871357

11. Chen, W., Zhao, H., & Li, Y. (2023). Mitochondrial dynamics in health and disease: mechanisms and potential targets. Signal Transduction and Targeted Therapy, 8(1), 333. 10.1038/s41392-023-01547-9

12. Cheng, Q., Liu, Q. Q., & Lu, C. (2024). A state-of-the-science review of using mitochondrial DNA copy number as a biomarker for environmental exposure. Environmental Pollution, 346, 123642. 10.1016/j.envpol.2024.123642

13. Clemente, D. B., Casas, M., Janssen, B. G., Lertxundi, A., Santa-Marina, L., Iñiguez, C., Llop, S., Sunyer, J., Guxens, M., Nawrot, T. S., & Vrijheid, M. (2017). Prenatal ambient air pollution exposure, infant growth and placental mitochondrial DNA content in the INMA birth cohort. Environmental Research, 157, 96–102. 10.1016/j.envres.2017.05.018

14. Clemente, D. B., Casas, M., Vilahur, N., Begiristain, H., Bustamante, M., Carsin, A., Fernández, M. F., Fierens, F., Gyselaers, W., Iñiguez, C., Janssen, B. G., Lefebvre, W., Llop, S., Olea, N., Pedersen, M., Pieters, N., Santa Marina, L., Souto, A., Tardón, A., . . . Nawrot, T. S. (2016). Prenatal ambient air pollution, placental mitochondrial DNA content, and birth weight in the INMA (Spain) and ENVIRONAGE (Belgium) birth cohorts. Environmental Health Perspectives, 124(5), 659–665. 10.1289/ehp.1408981

15. Cohen, A. J., Brauer, M., Burnett, R., Anderson, H. R., Frostad, J., Estep, K., Balakrishnan, K., Brunekreef, B., Dandona, L., Dandona, R., Feigin, V., Freedman, G., Hubbell, B., Jobling, A., Kan, H., Knibbs, L., Liu, Y., Martin, R., Morawska, L., . . . Forouzanfar, M. H. (2017). Estimates and 25-year trends of the global burden of disease attributable to ambient air pollution: an analysis of data from the Global Burden of Disease Study 2015. The Lancet, 389(10082), 1907–1918. 10.1016/s0140-6736(17)30505-6

16. DerSimonian, R., & Laird, N. (1986). Meta-analysis in clinical trials. Controlled Clinical Trials, 7(3), 177–188. 10.1016/0197-2456(86)90046-2

17. Ding, Y., Wan, Q., & Liu, W. (2025). Effects of atmospherically relevant PM2.5 on skeletal muscle mitochondria: a review of damage mechanisms and potential of exercise interventions. Frontiers in Public Health, 13, 1615363. 10.3389/fpubh.2025.1615363

18. Fuller, R., Landrigan, P. J., Balakrishnan, K., Bathan, G., Bose-O’Reilly, S., Brauer, M., Caravanos, J., Chiles, T., Cohen, A., Corra, L., Cropper, M., Ferraro, G., Hanna, J., Hanrahan, D., Hu, H., Hunter, D., Janata, G., Kupka, R., Lanphear, B., . . . Yan, C. (2022). Pollution and health: a progress update. The Lancet Planetary Health, 6(6), e535–e547. 10.1016/s2542-5196(22)00090-0

19. Gaikwad, A. S., Mahmood, R., Beerappa, R., Karunamoorthy, P., & Venugopal, D. (2020). Mitochondrial DNA copy number and cytogenetic damage among fuel filling station attendants. Environmental and Molecular Mutagenesis, 61(8), 820–829. 10.1002/em.22404

20. Grevendonk, L., Janssen, B. G., Vanpoucke, C., Lefebvre, W., Hoxha, M., Bollati, V., & Nawrot, T. S. (2016). Mitochondrial oxidative DNA damage and exposure to particulate air pollution in mother-newborn pairs. Environmental Health, 15(1), 10. 10.1186/s12940-016-0095-2

21. Gustafson, M. A., Sullivan, E. D., & Copeland, W. C. (2020). Consequences of compromised mitochondrial genome integrity. DNA Repair, 93, 102916. 10.1016/j.dnarep.2020.102916

22. Gustafsson, C. M., Falkenberg, M., & Larsson, N. (2016). Maintenance and expression of mammalian mitochondrial DNA. Annual Review of Biochemistry, 85(1), 133–160. 10.1146/annurev-biochem-060815-014402

23. Harrer, M., Cuijpers, P., Furukawa, T. A., & Ebert, D. D. (2021). Doing Meta-Analysis with R. 10.1201/9781003107347

24. Hautekiet, P., Nawrot, T. S., Janssen, B. G., Martens, D. S., De Clercq, E. M., Dadvand, P., Plusquin, M., Bijnens, E. M., & Saenen, N. D. (2020). Child buccal telomere length and mitochondrial DNA content as biomolecular markers of ageing in association with air pollution. Environment International, 147, 106332. 10.1016/j.envint.2020.106332

25. Hautekiet, P., Nawrot, T. S., Martens, D. S., Bijnens, E. M., De Keersmaecker, S. C., Van Der Heyden, J., De Clercq, E. M., & Saenen, N. D. (2025). Recent and chronic ambient air pollution exposure in association with telomere length and mitochondrial DNA content in the general population. Environmental Research, 276, 121525. 10.1016/j.envres.2025.121525

26. Hou, L., Zhang, X., Dioni, L., Barretta, F., Dou, C., Zheng, Y., Hoxha, M., Bertazzi, P. A., Schwartz, J., Wu, S., Wang, S., & Baccarelli, A. A. (2013). Inhalable particulate matter and mitochondrial DNA copy number in highly exposed individuals in Beijing, China: a repeated-measure study. Particle and Fibre Toxicology, 10(1), 17. 10.1186/1743-8977-10-17

27. Hou, L., Zhang, X., Wang, D., & Baccarelli, A. (2011). Environmental chemical exposures and human epigenetics. International Journal of Epidemiology, 41(1), 79–105. 10.1093/ije/dyr154

28. Hou, L., Zhu, Z., Zhang, X., Nordio, F., Bonzini, M., Schwartz, J., Hoxha, M., Dioni, L., Marinelli, B., Pegoraro, V., Apostoli, P., Bertazzi, P. A., & Baccarelli, A. (2010). Airborne particulate matter and mitochondrial damage: a cross-sectional study. Environmental Health, 9(1), 48. 10.1186/1476-069x-9-48

29. Hu, C., Qiao, J., Gui, S., Xu, K., Dzhambov, A. M., & Zhang, X. (2023). Perfluoroalkyl and polyfluoroalkyl substances and hypertensive disorders of pregnancy: A systematic review and meta-analysis. Environmental Research, 231(Pt 2), 116064. 10.1016/j.envres.2023.116064

30. Hu, C., Sheng, X., Li, Y., Xia, W., Zhang, B., Chen, X., Xing, Y., Li, X., Liu, H., Sun, X., & Xu, S. (2020). Effects of prenatal exposure to particulate air pollution on newborn mitochondrial DNA copy number. Chemosphere, 253, 126592. 10.1016/j.chemosphere.2020.126592

31. Janssen, B. G., Byun, H. M., Gyselaers, W., Lefebvre, W., Baccarelli, A. A., & Nawrot, T. S. (2015). Placental mitochondrial methylation and exposure to airborne particulate matter in the early life environment: An ENVIR*ON*AGE birth cohort study. Epigenetics, 10(6), 536–544. 10.1080/15592294.2015.1048412

32. Janssen, B. G., Munters, E., Pieters, N., Smeets, K., Cox, B., Cuypers, A., Fierens, F., Penders, J., Vangronsveld, J., Gyselaers, W., & Nawrot, T. S. (2012). Placental Mitochondrial DNA Content and Particulate Air Pollution during in Utero Life. Environmental Health Perspectives, 120(9), 1346–1352. 10.1289/ehp.1104458

33. Janssen, N. A., Hoek, G., Simic-Lawson, M., Fischer, P., van Bree, L., ten Brink, H., Keuken, M., Atkinson, R. W., Anderson, H. R., Brunekreef, B., & Cassee, F. R. (2011). Black carbon as an additional indicator of the adverse health effects of airborne particles compared with PM10 and PM2.5. Environmental health perspectives, 119(12), 1691–1699. 10.1289/ehp.1003369

34. Kaali, S., Jack, D., Delimini, R., Hu, L., Burkart, K., Opoku-Mensah, J., Quinn, A., Ae-Ngibise, K., Wylie, B., Boamah-Kaali, E., Chillrud, S., Owusu-Agyei, S., Kinney, P., Baccarelli, A., Asante, K., & Lee, A. G. (2018). Prenatal household air pollution alters cord blood mononuclear cell mitochondrial DNA Copy number: Sex-Specific Associations. International Journal of Environmental Research and Public Health, 16(1), 26. 10.3390/ijerph16010026

35. Kelly, F. J., & Fussell, J. C. (2015). Air pollution and public health: emerging hazards and improved understanding of risk. Environmental Geochemistry and Health, 37(4), 631–649. 10.1007/s10653-015-9720-1

36. Kim, K., Kabir, E., & Kabir, S. (2014). A review on the human health impact of airborne particulate matter. Environment International, 74, 136–143. 10.1016/j.envint.2014.10.005

37. Kwon, H., Ryu, M. H., & Carlsten, C. (2020). Ultrafine particles: unique physicochemical properties relevant to health and disease. Experimental & Molecular Medicine, 52(3), 318–328. 10.1038/s12276-020-0405-1

38. Lee, H., & Wei, Y. (2004). Mitochondrial biogenesis and mitochondrial DNA maintenance of mammalian cells under oxidative stress. The International Journal of Biochemistry & Cell Biology, 37(4), 822–834. 10.1016/j.biocel.2004.09.010

39. Li, R., Li, S., Pan, M., Chen, H., Liu, X., Chen, G., Chen, R., Yin, S., Hu, K., Mao, Z., Huo, W., Wang, X., Yu, S., Guo, Y., Hou, J., & Wang, C. (2021). Physical activity counteracted associations of exposure to mixture of air pollutants with mitochondrial DNA copy number among rural Chinese adults. Chemosphere, 272, 129907. 10.1016/j.chemosphere.2021.129907

40. Li, R., Zhou, R., & Zhang, J. (2018). Function of PM2.5 in the pathogenesis of lung cancer and chronic airway inflammatory diseases (Review). Oncology Letters, 15(5), 7506–7514. 10.3892/ol.2018.8355

41. Lin, C. H., Nicol, C. J. B., Wan, C., Chen, S. J., Huang, R. N., & Chiang, M. C. (2022). Exposure to PM_2.5_ induces neurotoxicity, mitochondrial dysfunction, oxidative stress and inflammation in human SH-SY5Y neuronal cells. Neurotoxicology, 88, 25–35. 10.1016/j.neuro.2021.10.009

42. Lin, C., Nicol, C. J., Wan, C., Chen, S., Huang, R., & Chiang, M. (2021). Exposure to PM2.5 induces neurotoxicity, mitochondrial dysfunction, oxidative stress and inflammation in human SH-SY5Y neuronal cells. NeuroToxicology, 88, 25–35. 10.1016/j.neuro.2021.10.009

43. Longchamps, R. J., Castellani, C. A., Yang, S. Y., Newcomb, C. E., Sumpter, J. A., Lane, J., Grove, M. L., Guallar, E., Pankratz, N., Taylor, K. D., Rotter, J. I., Boerwinkle, E., & Arking, D. E. (2020). Evaluation of mitochondrial DNA copy number estimation techniques. PLoS ONE, 15(1), e0228166. 10.1371/journal.pone.0228166

44. Mandakh, Y., Oudin, A., Erlandsson, L., Isaxon, C., Hansson, S. R., Broberg, K., & Malmqvist, E. (2021). Association of prenatal ambient air pollution exposure with placental mitochondrial DNA copy number, telomere length and preeclampsia. Frontiers in Toxicology, 3, 659407. 10.3389/ftox.2021.659407

45. Mishra, P. K., Bhargava, A., Kumari, R., Bunkar, N., Chauhan, P., Mukherjee, S., Shandilya, R., Singh, R. D., Tiwari, R., & Chaudhury, K. (2022). Integrated mitoepigenetic signalling mechanisms associated with airborne particulate matter exposure: A cross-sectional pilot study. Atmospheric Pollution Research, 13(5), 101399. 10.1016/j.apr.2022.101399

46. Mittal, M., Siddiqui, M. R., Tran, K., Reddy, S. P., & Malik, A. B. (2013). Reactive oxygen species in inflammation and tissue injury. Antioxidants and Redox Signaling, 20(7), 1126–1167. 10.1089/ars.2012.5149

47. Morris, R. H., Counsell, S. J., McGonnell, I. M., & Thornton, C. (2024). Exposure to urban particulate matter (UPM) impairs mitochondrial dynamics in BV2 cells, triggering a mitochondrial biogenesis response. The Journal of Physiology, 602(12), 2737–2750. 10.1113/jp285978

48. Murphy, M. P., & Hartley, R. C. (2018). Mitochondria as a therapeutic target for common pathologies. Nature Reviews Drug Discovery, 17(12), 865–886. 10.1038/nrd.2018.174

49. Ohlwein, S., Kappeler, R., Kutlar Joss, M., Künzli, N., & Hoffmann, B. (2019). Health effects of ultrafine particles: a systematic literature review update of epidemiological evidence. International Journal of Public Health, 64(4), 547–559. 10.1007/s00038-019-01202-7

50. Peng, C., Cayir, A., Sanchez-Guerra, M., Di, Q., Wilson, A., Zhong, J., Kosheleva, A., Trevisi, L., Colicino, E., Brennan, K., Dereix, A. E., Dai, L., Coull, B. A., Vokonas, P., Schwartz, J., & Baccarelli, A. A. (2017). Associations of Annual Ambient Fine Particulate Matter Mass and Components with Mitochondrial DNA Abundance. Epidemiology, 28(6), 763–770. 10.1097/ede.0000000000000717

51. Picard, M., & Shirihai, O. S. (2022). Mitochondrial signal transduction. Cell Metabolism, 34(11), 1620– 1653. 10.1016/j.cmet.2022.10.008

52. Pieters, N., Koppen, G., Smeets, K., Napierska, D., Plusquin, M., De Prins, S., Van De Weghe, H., Nelen, V., Cox, B., Cuypers, A., Hoet, P., Schoeters, G., & Nawrot, T. S. (2013). Decreased Mitochondrial DNA Content in Association with Exposure to Polycyclic Aromatic Hydrocarbons in House Dust during Wintertime: From a Population Enquiry to Cell Culture. PLoS ONE, 8(5), e63208. 10.1371/journal.pone.0063208

53. Qiao, J., Sun, L., Zhang, M., Gui, S., Wang, X., & Hu, C. (2024). Association between ambient particulate matter exposure and mitochondrial DNA copy number: A systematic review and meta-analysis. The Science of the Total Environment, 923, 171423. 10.1016/j.scitotenv.2024.171423

54. Qiu, Y., Huang, Y., Chen, M., Yang, Y., Li, X., & Zhang, W. (2022). Mitochondrial DNA in NLRP3 inflammasome activation. International immunopharmacology, 108, 108719. 10.1016/j.intimp.2022.108719

55. Rosa, M. J., Just, A. C., Guerra, M. S., Kloog, I., Hsu, H. L., Brennan, K. J., García, A. M., Coull, B., Wright, R. J., Rojo, M. M. T., Baccarelli, A. A., & Wright, R. O. (2016). Identifying sensitive windows for prenatal particulate air pollution exposure and mitochondrial DNA content in cord blood. Environment International, 98, 198–203. 10.1016/j.envint.2016.11.007

56. Schraufnagel, D. E., Balmes, J. R., Cowl, C. T., De Matteis, S., Jung, S., Mortimer, K., Perez-Padilla, R., Rice, M. B., Riojas-Rodriguez, H., Sood, A., Thurston, G. D., To, T., Vanker, A., & Wuebbles, D. J. (2018). Air pollution and noncommunicable diseases. CHEST Journal, 155(2), 409–416. 10.1016/j.chest.2018.10.042

57. Sivakumar, B., & Kurian, G. A. (2024). Investigating the temporal link between PM2.5 exposure and acceleration of myocardial ischemia reperfusion injury: Emphasizing the hazardous presence of metals in inhaled air. Environmental Pollution, 355, 124113. 10.1016/j.envpol.2024.124113

58. Smith, A. R., Hinojosa Briseño, A., Picard, M., & Cardenas, A. (2023). The prenatal environment and its influence on maternal and child mitochondrial DNA copy number and methylation: A review of the literature. Environmental research, 227, 115798. 10.1016/j.envres.2023.115798

59. Stoccoro, A., & Coppedè, F. (2021). Mitochondrial DNA methylation and human diseases. International Journal of Molecular Sciences, 22(9), 4594. 10.3390/ijms22094594

60. Thompson, R., Smith, R. B., Bou Karim, Y., Shen, C., Drummond, K., Teng, C., & Toledano, M. B. (2022). Noise pollution and human cognition: An updated systematic review and meta-analysis of recent evidence. Environment international, 158, 106905. 10.1016/j.envint.2021.106905

61. Van Der Stukken, C., Nawrot, T. S., Wang, C., Lefebvre, W., Vanpoucke, C., Plusquin, M., Roels, H. A., Janssen, B. G., & Martens, D. S. (2022). The association between ambient particulate matter exposure and the telomere–mitochondrial axis of aging in newborns. Environment International, 171, 107695. 10.1016/j.envint.2022.107695

62. Vos, S., Nawrot, T. S., Martens, D. S., Byun, H., & Janssen, B. G. (2020). Mitochondrial DNA methylation in placental tissue: a proof of concept study by means of prenatal environmental stressors. Epigenetics, 16(2), 121–131. 10.1080/15592294.2020.1790923

63. Vrijheid, M., Martinez, D., Manzanares, S., Dadvand, P., Schembari, A., Rankin, J., & Nieuwenhuijsen, M. (2010). Ambient Air pollution and Risk of congenital Anomalies: A Systematic review and Meta-analysis. Environmental Health Perspectives, 119(5), 598–606. 10.1289/ehp.1002946

64. Wang, X., Hart, J. E., Liu, Q., Wu, S., Nan, H., & Laden, F. (2020). Corrigendum to “Association of particulate matter air pollution with leukocyte mitochondrial DNA copy number” [Environ. Int. 141 (2020) 105761]. Environment International, 143, 106069. 10.1016/j.envint.2020.106069

65. Whaley, P., Aiassa, E., Beausoleil, C., Beronius, A., Bilotta, G., Boobis, A., de Vries, R., Hanberg, A., Hoffmann, S., Hunt, N., Kwiatkowski, C. F., Lam, J., Lipworth, S., Martin, O., Randall, N., Rhomberg, L., Rooney, A. A., Schünemann, H. J., Wikoff, D., Wolffe, T., … Halsall, C. (2020). Recommendations for the conduct of systematic reviews in toxicology and environmental health research (COSTER). Environment international, 143, 105926. 10.1016/j.envint.2020.105926

66. Wong, J. Y., Hu, W., Downward, G. S., Seow, W. J., Bassig, B. A., Ji, B., Wei, F., Wu, G., Li, J., He, J., Liu, C., Cheng, W., Huang, Y., Yang, K., Chen, Y., Rothman, N., Vermeulen, R. C., & Lan, Q. (2017). Personal exposure to fine particulate matter and benzo[a]pyrene from indoor air pollution and leukocyte mitochondrial DNA copy number in rural China. Carcinogenesis, 38(9), 893–899. 10.1093/carcin/bgx068

67. Xu, W., Xu, X., Zhang, Y., Li, F., & Xia, D. (2025). Mitochondrial fission and fusion in inflammatory diseases: mechanisms and therapeutic implications. Journal of Translational Medicine, 24(1), 127. 10.1186/s12967-025-07605-w

68. Yin, M., & Guo, L. (2025). Mitochondrial DNA methylation: State-of-the-art in molecular mechanisms and disease implications. Journal of Advanced Research, 83, 455–473. 10.1016/j.jare.2025.08.029

69. Zhang, G., Jiang, F., Chen, Q., Yang, H., Zhou, N., Sun, L., Zou, P., Yang, W., Cao, J., Zhou, Z., & Ao, L. (2020). Associations of ambient air pollutant exposure with seminal plasma MDA, sperm mtDNA copy number, and mtDNA integrity. Environment International, 136, 105483. 10.1016/j.envint.2020.105483

70. Zhang, Y., Dong, S., Wang, H., Tao, S., & Kiyama, R. (2016). Biological impact of environmental polycyclic aromatic hydrocarbons (ePAHs) as endocrine disruptors. Environmental Pollution, 213, 809–824. 10.1016/j.envpol.2016.03.050

71. Zhong, J., Cayir, A., Trevisi, L., Sanchez-Guerra, M., Lin, X., Peng, C., Bind, M., Prada, D., Laue, H., Brennan, K. J., Dereix, A., Sparrow, D., Vokonas, P., Schwartz, J., & Baccarelli, A. A. (2015). Traffic-Related air pollution, blood pressure, and adaptive response of mitochondrial abundance. Circulation, 133(4), 378–387. 10.1161/circulationaha.115.018802

72. Zong, Y., Li, H., Liao, P., Chen, L., Pan, Y., Zheng, Y., Zhang, C., Liu, D., Zheng, M., & Gao, J. (2024). Mitochondrial dysfunction: mechanisms and advances in therapy. Signal Transduction and Targeted Therapy, 9(1), 124. 10.1038/s41392-024-01839-8

