## Supplementary for "Effect of Airborne Particulate Matter on mtDNA Copy Number: A Systematic Review and Meta- Analysis": Supplementary Information.docx

**Supplementary information showing “Search Strategy” for databases**

**PubMed 320 articles**

("Air Pollution"[Mesh] OR "Particulate Matter"[Mesh] OR

"Air pollution" OR "particulate matter" OR PM2.5 OR PM10 OR

"Ultrafine particles" OR "diesel exhaust particles" OR "black carbon")

AND

("DNA, Mitochondrial"[Mesh] OR

"mtDNA copy number" OR "mitochondrial DNA methylation" OR

"Mito epigenetic" OR "mitochondrial dysfunction" OR

"Mitochondrial gene expression")

NOT

("Review"[Publication Type] OR

"Systematic Review"[Publication Type] OR

"Meta-Analysis"[Publication Type] OR

"Editorial"[Publication Type] OR

"Letter"[Publication Type] OR

"Comment"[Publication Type])

**SCOPUS 214 articles**

("air pollution" OR "particulate matter" OR PM2.5 OR PM10)

AND

("mitochondrial DNA" OR "mtDNA copy number" OR "mtDNA methylation")

AND

("mitochondrial dysfunction" OR "mitochondrial gene expression")

**SCOPUS 58 ARTICLES**

airborne particulate matter' AND 'mitochondrial DNA copy number

**SCOPUS 8 articles**

("PM2.5" OR "PM10")

AND

("mtDNA copy number" OR "mitochondrial DNA methylation")

AND

("human study" OR "animal experiment")

**Web of Science**

("PM2.5" OR "PM10" OR "particulate matter")

AND

("mitochondrial DNA copy number" OR "mtDNA methylation")

AND

("human study" OR "animal experiment")

**Keywords**

Particulate matter; Mitochondrial DNA; Epigenetic inheritance; Air pollution

**Boolean operators**

(AND, OR, and NOT)
